# Mathematical modelling of SARS-CoV-2 variant outbreaks reveals their probability of extinction

**DOI:** 10.1101/2021.07.05.21260005

**Authors:** Henrik Schiøler, Torben Knudsen, Rasmus Froberg Brøndum, Jakob Stoustrup, Martin Bøgsted

## Abstract

When a virus spreads, it may mutate into, e.g., vaccine resistant or fast spreading lineages, as was the case for the Danish Cluster-5 mink variant (belonging to the B.1.1.298 lineage), the British B.1.1.7 lineage, and the South African B.1.351 lineage of the SARS-CoV-2 virus. A way to handle such spreads is through a containment strategy, where the population in the affected area is isolated until the spread has been stopped. Under such circumstances, it is important to monitor whether the mutated virus is extinct via massive testing for the virus sub-type. If successful, the strategy will lead to lower and lower numbers of the sub-type, and it will eventually die out. An important question is, for how long time one should wait to be sure the sub-type is extinct? We use a hidden Markov model for infection spread and an approximation of a two stage sampling scheme to infer the probability of extinction. The potential of the method is illustrated via a simulation study. Finally, the model is used to assess the Danish containment strategy when SARS-CoV-2 spread from mink to man during the summer of 2020, including the Cluster-5 sub-type. In order to avoid further spread and mink being a large animal virus reservoir, this situation led to the isolation of seven municipalities in the Northern part of the country, the culling of the entire Danish 17 million large mink population, and a bill to interim ban Danish mink production until the end of 2021.

## 1 Introduction

Pandemic outbreaks have reentered as a global reality and threat to humanity with the transmission of an animal-adapted Corona virus to humans, first detected in Wuhan, China in late 2019, leading to the COVID-19 pandemic exhibiting frequent severe respiratory problems in humans. Early warnings of a global event were seen with SARS and avian flu [3, 9]. In both cases early containment measures proved successful, whereas for SARS-CoV-2 early containment failed and the strategy transferred to mitigation. This pattern has later been re-observed in almost all countries at the early stages of COVID-19 introduction across national borders. Lately, human-animal transmission has given rise to grave concerns regarding a re-ignition of the pandemic through resistant mutations cultivated in animal reservoirs [11]. One such example is the discovery of the Cluster-5 variant in humans transferred from farmed mink in the Danish fur industry during the summer of 2020 [2]. Cluster-5 belongs to the B.1.1.298 lineage and is characterized by 11 amino acid substitutions and four amino acid deletions relative to Wuhan-Hu-1. It was indicated that Cluster-5 could be vaccine resistant [8]. Hence, National and global health concerns triggered severe disease containment measures, such as the rapid culling of the entire Danish 17 million large stock of mink as well as relatively severe social- and travel-restrictions for seven municipalities in the North Denmark Region (approx. 281,000 people). Containment measures were, for various reasons, delayed for around four weeks, in which there were no observations of Cluster-5 mutations in a subset of polymerase chain reaction (PCR) tested samples subjected to whole genome sequencing (WGS). This has lead to the primary objective of the present paper, namely to answer the question: For how long should Cluster-5 be absent from test samples before its extinction is sufficiently certain? The answer depends on the epidemiological behaviour of the disease during restrictions as well as the testing regime imposed in that period. We aim in this paper to provide a Bayesian model-based answer to this question which links epidemiological parameters as well as testing patterns and test results to the probability of disease extinction and early detection.

Various modeling levels exist in epidemiology such as compartment models, aggregate Markov models, and individual Markov models [1]. Whereas the former two, including the well known SIR and SEIR models [6], are well suited to model the epidemic spread for large populations during mitigation or endemic spread if the virus spread reaches steady state [4], the latter provides higher precision for small amounts of infected during containment. Moreover, the primary objective of the paper is not possible to pursue with deterministic models. Other recent investigations have been made to model the early epidemic evolution of SARS-CoV-2, employing auto-regressive modeling with a Bayesian approach to parameter estimation [10]. Such models provide mean value predictions but do not give the probabilistic output as requested above. The scale of genomic surveillance needed for early detection of newly emerging variants of concern (VoC) has been considered through a model of the sampling process including the PCR test quality parameters [12]. However, in this model, only the output model is considered, in contrast to our model, where also the epidemic dynamics are included. Furthermore, results are given as expected counts in contrast to the probabilistic results of our approach. A generalized *Hidden* Markovian model framework for epidemic evolution and test has also been employed [13]. One may consider the model class used in this paper as a subset of that model, tailored specifically to early epidemic development, which brings about a much required computational tractability even for large populations.

We shall shortly introduce the development from individual models to compartment models to facilitate the transfer of model parameters between them. The model is generic and can therefore be used in other situations when pathogen mutations are entered from, e.g., animal reservoirs.

## 2 Results

The derivation of the epidemic spread and measurement model was motivated by the spread of mink mutations in the North Denmark Region. Before returning to this, we will formulate the model and study its usability and robustness by running a number of intervention scenarios. In the following we will consider interventions as a combination of restrictions, bringing the reproduction number down, and intensified PCR and WGS sequencing.

### 2.1 Probability of extinction

Assume a situation where we have observed *y* infected people carrying a variant we wish to keep under control and an effective containment strategy of infected people and their immediate contacts has been invoked. The question is now:, for how long shall we retain the restrictions to be reasonably sure that the virus has not spread? I.e., we want to calculate the following probability

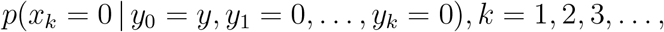

where *x*_*k*_ and *y*_*k*_ are, respectively, the hidden (true) and observed number of infected people carrying the variant at time *k*.

In the Methods section, we have formulated a discrete time hidden Markov model to model this situation where the development of the number of infected people, with the specific variant of interest, follows a birth-death process with death rate (herein recovery rate) *γ* and net reproduction rate *R*_0_. The net reproduction rate is defined as the ratio of the birth rate (herein infection rate) versus the death rate, i.e., *R*_0_ = *β/γ*. We assume a two-step testing strategy where *n*_*k*_ of the population of size *N*, are PCR tested and *m*_*k*_ of the PCR positive tests are WGS tested at time point *k*.

In the following, we compute a number of scenarios which illustrate how various intervention strategies will influence the time until a certain probability of extinction has been reached, given the specific variant has not been observed for a given period of time. In all simulations, we assume a constant recovery time of two weeks, i.e. *γ* = 0.5, a population size of *N* = 600, 000, *n* = 10, 000 tests per week, and an initial number of infected people with the specific variant of 11 as well as a flat prior distribution on the number of specific cases. These numbers were picked to mimic the Cluster-5 outbreak in the North Denmark Region, where 11 cases were observed in a population of size approximately 600,000. Thereafter, we simulated increased restrictions by lowering stepwise the reproduction rate, *R*_0_, from 1.5 to 0.5. Finally, we studied increased WGS testing rates of positives between 1% and 75%.

In Figure 1, Panel A shows the probability of extinction as a function of the number of weeks for increasing WGS ratio and a constant reproduction rate of *R*_0_ = 1.0, and Panel B shows the probability of extinction as function of the number of weeks for increasing reproduction rates and constant WGS rate of 0.25. Time to the probability of extinction for all scenarios can be seen in Table 1.

**Table 1:** Weeks to thresholds for various testing and restriction strategies.

| WGS ratio | $R_0$ | Prob < 0.85 | Prob < 0.90 | Prob < 0.95 |
| --- | --- | --- | --- | --- |
| 0.01 | 0.50 |  |  |  |
|  | 0.75 |  |  |  |
|  | 1.00 |  |  |  |
|  | 1.25 | 26 | 30 | 35 |
|  | 1.50 | 16 | 18 | 21 |
| 0.25 | 0.50 | 39 | 40 | 42 |
|  | 0.75 | 41 | 44 | 49 |
|  | 1.00 | 41 | 46 | 54 |
|  | 1.25 | 23 | 26 | 30 |
|  | 1.50 | 15 | 17 | 20 |
| 0.5 | 0.50 | 28 | 30 | 32 |
|  | 0.75 | 34 | 36 | 40 |
|  | 1.00 | 32 | 35 | 41 |
|  | 1.25 | 21 | 23 | 27 |
|  | 1.50 | 15 | 16 | 19 |
| 0.75 | 0.50 | 26 | 27 | 30 |
|  | 0.75 | 30 | 32 | 36 |
|  | 1.00 | 27 | 30 | 35 |
|  | 1.25 | 20 | 22 | 25 |
|  | 1.50 | 14 | 16 | 18 |

**Figure 1:**
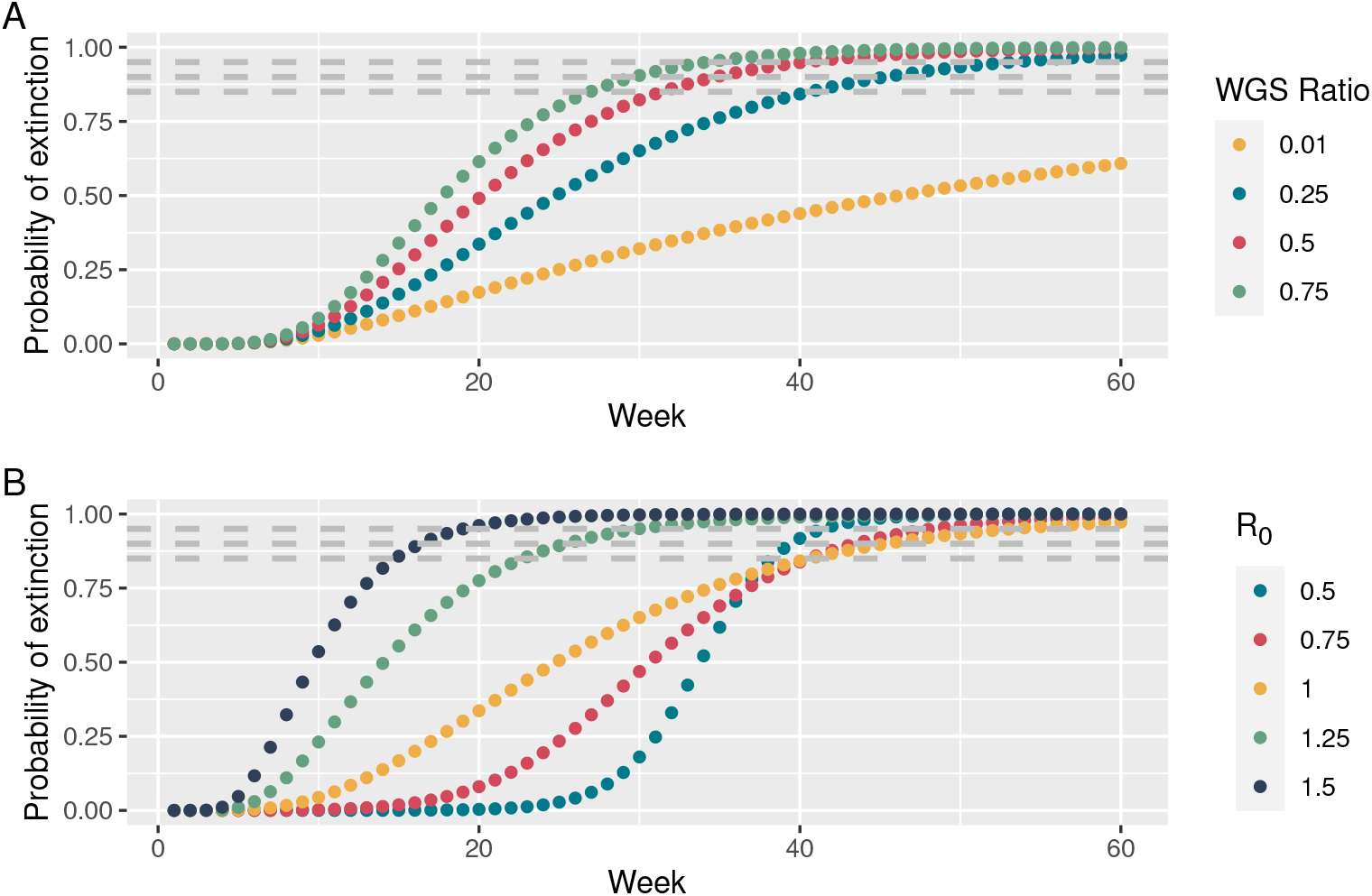
Probability of extinction as a function of the number of weeks with no observations of the specific variant. Dashed lines indicate probability levels 0.85, 0.90, and 0.95, respectively. (A) Variation of testing regime for a constant reproduction rate of *R*_0_ = 1. (B) Variation of restrictions (*R*_0_) for a constant WGS testing ratio of 0.25. The yellow curves are identical in the two plots.

From numerical results, we see that an increase in the ratio of WGS tests dramatically lowers the number of weeks from 42 to 25 before we can conclude a probability of extinction of 90%. We also noticed a counter-intuitive non-monotone relationship between reproduction rate and number of weeks until a certain probability of extinction has been achieved. To investigate this further, we computed the number of weeks to a 85%, 90%, and 95% probability of extinction and depicted the number of weeks to extinction against increasing reproduction rates, ranging from 0.5 to 2.5, see Figure 2. From this we notice the maximum of weeks to probability of extinction emerging for reproduction rates *R*_0_ slightly less than one, and decreasing for higher values. We acknowledge the counter-intuitive behaviour that weeks-to-extinction decreases as *R*_0_ increases. The behavior can intuitively be explained by the argument that if the epidemic has a high growth rate, it is unrealistic, if it is present, that it has not been seen. We can also attribute this effect to the often-experienced counter-intuitive nature of a-posterior probabilities, where a-priori probabilities may decrease whereas conditional observation probabilities may increase altogether yielding a non-monotonous a-posterior probability. More specifically, under fast epidemic growth the lacking observation of positive cases may contribute higher to the a-posterior probability of extinction than it would under a slower growth or decay. I.e., under higher growth, it is more surprising to observe zero positives than under lower growth or decay. In other words, if, under fast growth, the variant is not extinct, it would be highly unlikely to observe zero positives. This may reflect back onto disease control, since tightened restrictions may indeed prolong the period until probable extinction even when decreasing reproduction number across the *R*_0_ = 1 boundary. Of course, any reduction in *R*_0_ would increase extinction likelihood (a-priori probability), but decision makers need to be aware of the former effect, when deciding to end, e.g., a lock down period. We have in the Supplementary Information, provided a simple example showing the same counter-intuitive effect.

**Figure 2:**
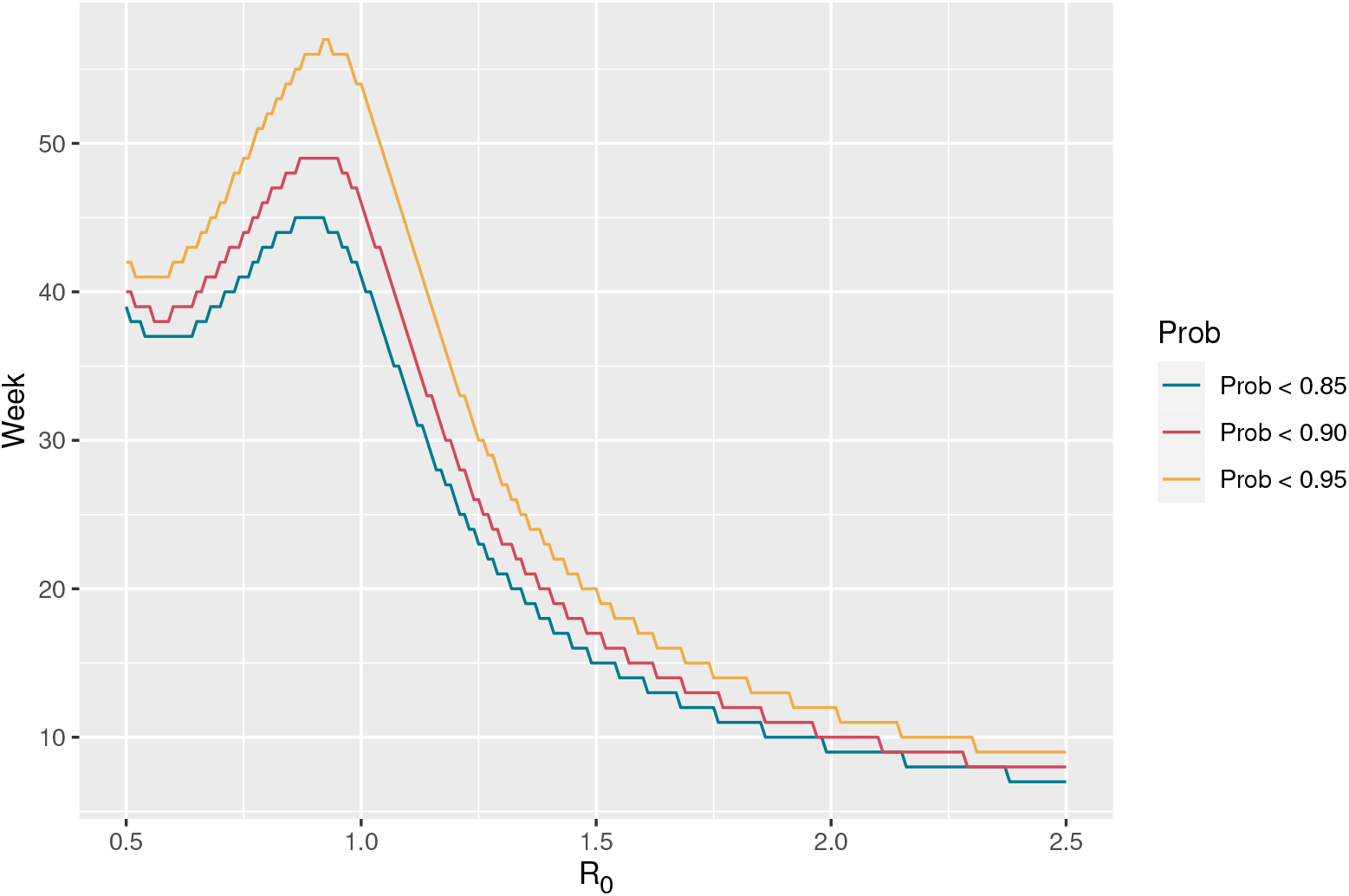
Weeks to extinction as function of the reproduction rate, *R*_0_.

We are aware that it is impossible to set all parameters for a given situation. We have therefore made an online Shiny App which can be used to compute the interested reader’s own scenarios, please refer to the Code availability section.

### 2.2 Analysis of the Cluster-5 extinction in Denmark

Denmark has a total population of 5.8 million and is divided into 98 municipalities which are organized in five administrative regions. The North Denmark Region has 590,000 inhabitants and contains the 11 most northern municipalities, see the coloured municipalities in Figure 3. All population statistics are from December 31, 2020 and have been fetched from the Statbank of Statistics Denmark. For further details see the Data availability section.

**Figure 3:**
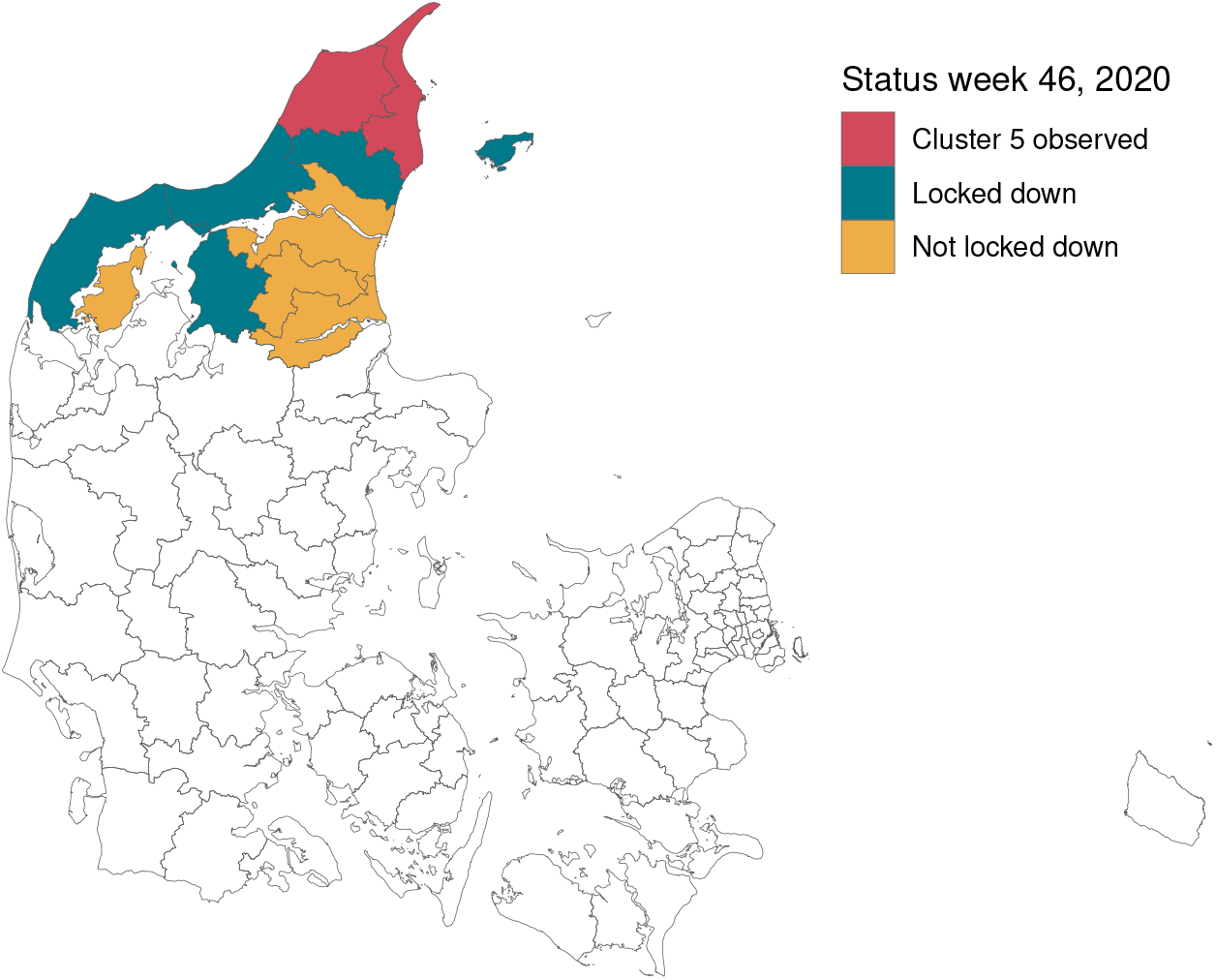
Status for the municipalities in the North Denmark Region following the lock-down order on November 6, 2020. The order was imposed in municipalities with observations of Cluster-5 (red), as well as surrounding municipalities with a high concentration of mink farms (blue). The remaining coloured municipalities belong to the North Denmark Region, but was not locked down. The remaining municipalities in Denmark are coloured white.

The Cluster-5 variant was only observed in the two most northern municipalities Hjørring and Frederikshavn, shown in red in Figure 3. The Figure also shows the seven municipalities covered by the Danish Government’s lock-down (in red and blue), amounting to 281,000 inhabitants.

During a period of 4 weeks from mid August 2020 to mid September 2020 (week no. 35-38), respectively 3, 3, 1, and 4, Cluster-5 observations were made. The public was warned by the authorities against the potential vaccine resistant Cluster-5 variant on November 6, 2020, and it was decided by the authorities to cull the entire 17 million large Danish mink population and lock-down seven municipalities in the North Denmark Region to hinder further spread of the variant. The lock-down was planned to run from November 9, 2020, till December 7, 2020, i.e. week 46 to 49. However, due to low infection rates and heavy political pressure the strict restrictions were removed after only two weeks, i.e. at the beginning of week 47. One of the persisting questions from the Danish press and political opposition has been whether Cluster-5 was extinct with a reasonably high probability. In the following, we will try to shed light on this question.

We compare the two situations: planned and actually realized, which apart from the shortened period of intervention mainly differs in the number of WGS tests actually conducted. From publicly available data, we could only get access to week-by-week summary statistics for the entire North Denmark Region. Data from the Cluster-5 outbreak until the end of 2020 can be seen in Table 2. The Data have been obtained from the official Danish Epidemiological Report, for further details see the Data availability section.

**Table 2:** Weekly test data.

| Week | PCR | Infected | WGS | Pct. WGS | Cluster5 |
| --- | --- | --- | --- | --- | --- |
| 35 | 22539 | 49 | 25 | 51 | 3 |
| 36 | 23758 | 67 | 27 | 40 | 3 |
| 37 | 29823 | 233 | 43 | 18 | 1 |
| 38 | 40681 | 331 | 74 | 22 | 4 |
| 39 | 39789 | 292 | 80 | 27 | 0 |
| 40 | 28621 | 260 | 40 | 15 | 0 |
| 41 | 29177 | 295 | 47 | 16 | 0 |
| 42 | 23125 | 329 | 138 | 42 | 0 |
| 43 | 36456 | 679 | 39 | 6 | 0 |
| 44 | 52099 | 819 | 177 | 22 | 0 |
| 45 | 64493 | 588 | 216 | 37 | 0 |
| 46 | 93577 | 485 | 308 | 64 | 0 |
| 47 | 95742 | 357 | 246 | 69 | 0 |
| 48 | 46120 | 267 | 170 | 64 | 0 |
| 49 | 33153 | 387 | 64 | 17 | 0 |
| 50 | 51438 | 964 | 104 | 11 | 0 |
| 51 | 81265 | 1422 | 344 | 24 | 0 |
| 52 | 61935 | 1303 | 362 | 28 | 0 |
| 53 | 57121 | 1222 | 270 | 22 | 0 |

We used a population size of 281,000 and divided the number of PCR tests, WGS tests and positives by two, as the locked-downed municipalities correspond to approx. half of the population. Further we set the recovery rate to 0.5 (i.e., two weeks) and the reproduction number before intervention to 1.2.

During intervention, the plan was to test the entire population of the municipalities over a 4-week period as well as WGS testing all positive samples.The number of PCR tests were therefore set to 281,000/4 = 70250 PCR tests per week. If we assume a positive pct. of 1.5%, we get 1100 positive tests. The test capacity was up to 5000 a week, so we assume all 1100 would be WGS tested during intervention according to the plan. These assumptions are off-course too optimistic as the viral load and quality can be too low for sequencing. However, this could be seen as an upper limit on the performance. The reproduction number of the new variant can either be worse, neutral, or improved compared to the original variant. In this example, we assume that the net effects of a changed reproduction number and lock-down leads to a reproduction rate of 1.0 during lock-down.

The week-to-week assessment of the probability of extinction from the Cluster-5 outbreak in week 35 till the planned lock-down is depicted in Figure 4. One sees how the probability of extinction develops under the planned intervention strategy and what was realized. We see the probability was 0.22 before the intervention and 0.37 when the restrictions were lifted and 0.52 if the restrictions had been lifted December 3, 2020.

**Figure 4:**
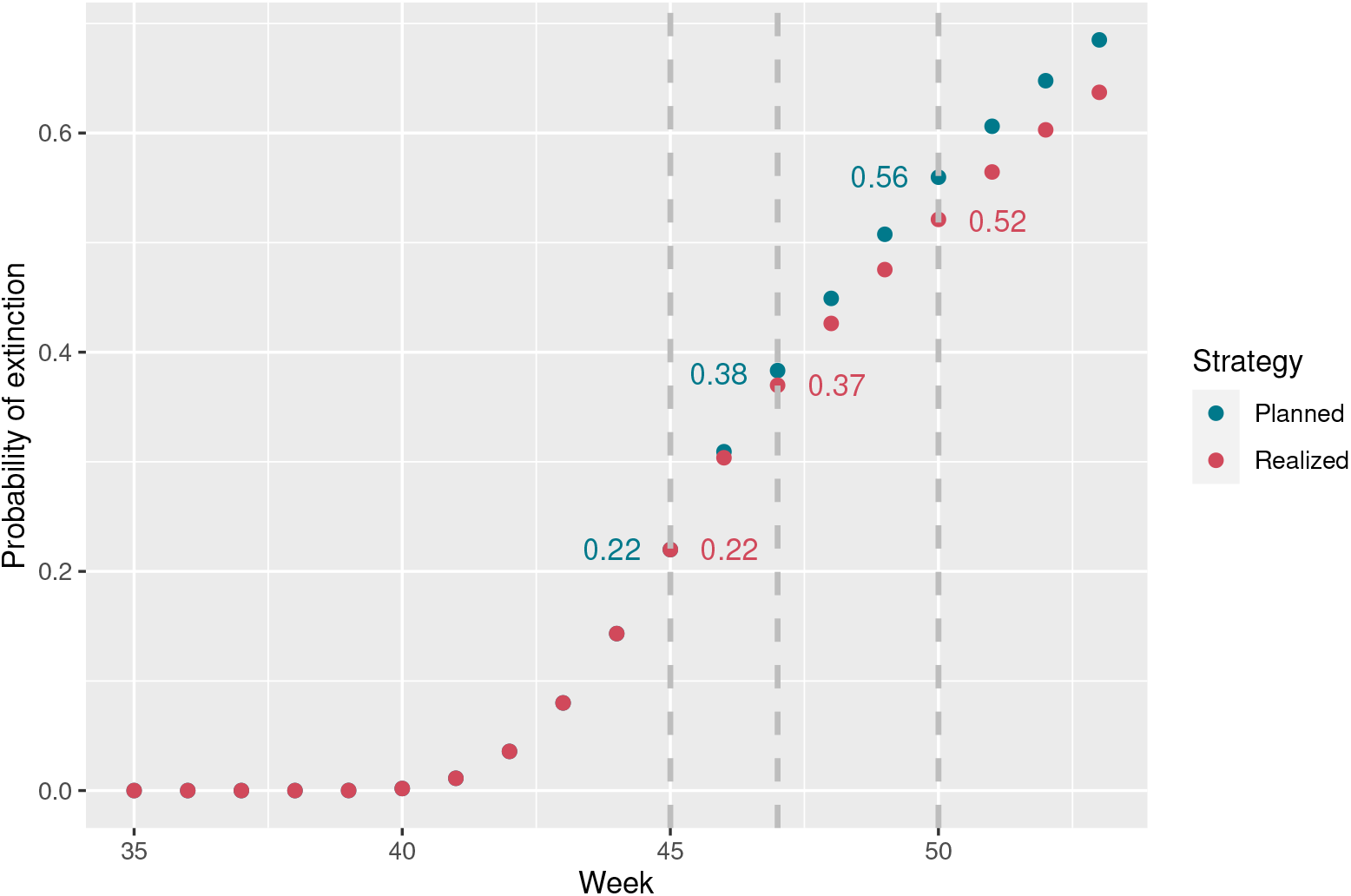
Probability of extinction for planned and realized interventions.

## 3 Discussion

Using Bayes filtering of a hidden Markov model with realistic parameters based on the Cluster-5 variant case from Denmark, we were able to quantify the impact of interventions on the certainty of extinction of deleterious SARS-CoV-2 variants. We found counter-intuitively that imposing restrictions in general increases the time to certainty of variant extinction, wherefore restrictions should be supplemented by a massive testing strategy. For the Danish case, we concluded a low probability of extinction when the restrictions were lifted at the beginning of week 46. However, at the time of writing (March 1, 2021), the variant has not emerged, so the probability of extinction is now well above 0.95%. However, one should be aware that the calculations are based on rough estimates. The calculations could be made much more exact, if we have had access to the detailed recordings from the Danish authorities.

Although, the use of birth-death processes to model the extinction of species is not new, we have not been able to find previous research with an attempt to calculate the probability of extinction based on hidden information about the birth-death process [7].

The work provides a simple and fast computational framework. This implies a number of scenarios, including sensitivity analyses that can quickly be computed. The simplified model used here is ideal for the initial outbreak of a new variant of concern, whereas other model frameworks such as compartment model (SIR, SEIR) are more well suited later in the epidemic evolution, i.e. when some variant is wider spread.

In conclusion, we hope this tools will be useful for decision makers when deciding upon intervention strategies, that effectively balance restrictions and test strategies.

## 4 Methods

In order to formulate the hidden Markov model, we use the notation in Box 1:

**Box 1: Mathematical notation**

- Population characteristics
  – *N* population size
  – *X*_*k*_ number of infected with the specific variant
  – 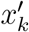 number of infected with a non-specific variant
  – 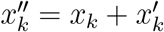 total number of infected
- PCR test statistics
  – *n*_*k*_ sample size of the PCR test
  – *z*_*k*_ number of samples with the specific variant
  – 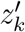 number of samples with a non-specific variant
- WGS test statistics
  – *m*_*k*_ sample size of the WGS test (*≤ n*_*k*_)
  – 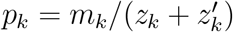 ratio of WGS tested out of positive PCR tests
  – *y*_*k*_ number of samples with the specific variant
- Epidemic parameters
  – *β* infection rate
  – *γ* recovery rate
  – *R*_*0*_ = *β /γ* net reproduction rate

### 4.1 The epidemic model

We first consider the elementary infection dynamics between two persons P1 and P2 of which P1 is infected and P2 is susceptible. Consider an infinitesimal time interval [*t, t* + *dt*] where P1 and P2 are within infection range. Modelling the infection state of P2 as a two-state Continuous Time Markov Chain (CTMC), yields the probability of P1 infecting P2 within [*t, t* + *dt*], to be *b dt*, where *b* is a disease characteristic constant.

Consider then a susceptible individual P interacting with a population, comprising *x* infected of a population size *N* If it is assumed that P on the average finds *L* others within his/her range of infection then the average probability of P being infected within [*t,t* + *dt*] is 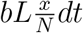.

Consider next *S* susceptible individuals each interacting with a population, comprising *x* infected of a population size *N*. Then the probability of 1 out of the susceptible individuals being infected in [*t, t* + *dt*] is approximately

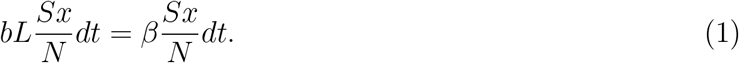

This leads to the following differential equation governing the evolution of expectations

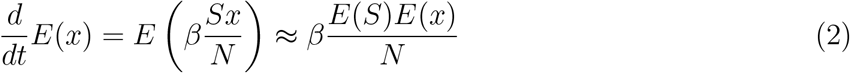

comprising the infection rate equation of the SIR model. When an *exposed* state is inserted between susceptible and infected states, (2) would yield the rate of transfers between susceptible and exposed states.

Considering instead of expectations, a probability distribution over the actual number of infected *I*, (1) leads to

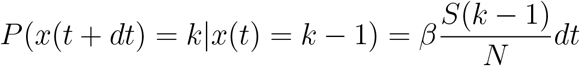

and with the Bayes law of total probability

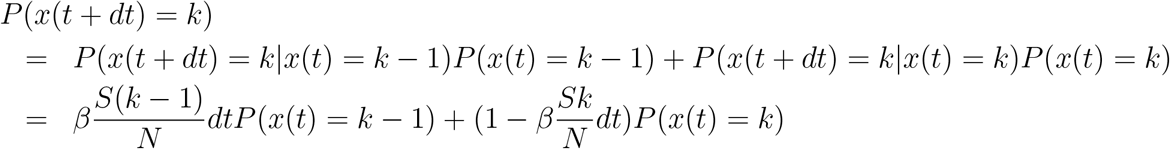

yielding

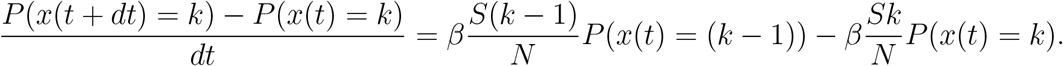

Leading to the differential equation

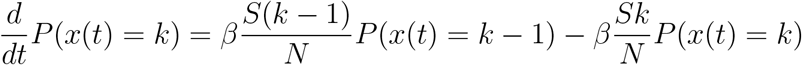

For the early development of an outbreak *S ≈ N*. This yields the infection dynamics

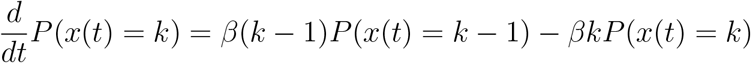

Adding the effect of recovery, we obtain

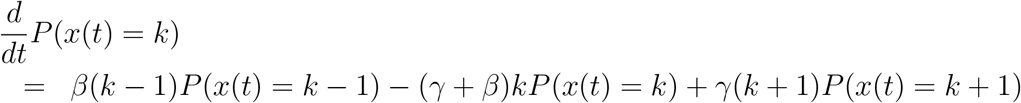

where *γ* is the individual recovery rate. This can altogether be summarized by the CTMC depicted in Figure 5.

**Figure 5:**
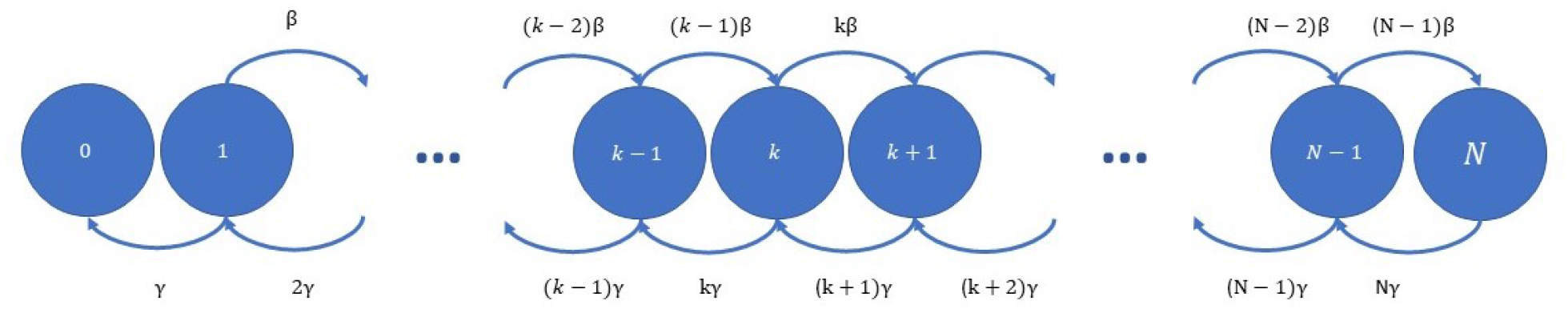
Continuous Time Markov Chain for Cluster-5 infected.

Thus, the number of infected people under the epidemic can be modelled as a continuous time Markov chain (CTMC) *{x*_*t*_, *t ≥* 0*}*, with state space *X* = *{*0, 1, 2, …, *N }* and infinitesimal generator *Q*, where Q is a matrix with elements, for *i, j ∈ {*1, 2, 3, …, *N }*,

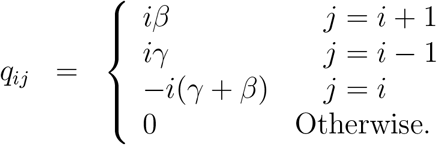

We can now model the daily number of infected in the population as a discretely sampled CTMC *{x*(*n*), *n* = 0, 1, 2, … *}*, with state space *X* = *{*0, 1, 2, …, *N }* and transition probabilities

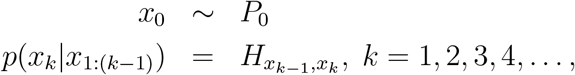

where *P*_0_ = (*p*(*x*_0_ = 0), *p*(*x*_0_ = 1), …, *p*(*x*_0_ = *N*)) is the initial distribution of *x*_0_ and 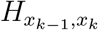 is the *x*_*k* − 1_, *x*_*k*_ ‘th element of the matrix *H* = exp(*Q dT*), with *dT* being the sampling period.

The presence of the transmitted virus among humans is first detected through an initial sample of test results *y*_0_. Therefore the initial conditions for the Bayes filter may be found from

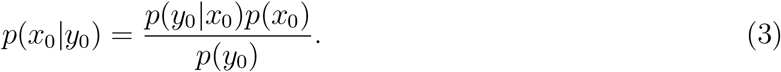

In most cases, if there is no initial evidence for *x*_0_, then we may (using the principle of maximum entropy) a priory assume *x*_0_ is uniformly distributed over some interval, e.g, *{*0,.., *N }* (coined uniform in the accompanying R-script). Another possibility is to choose *x*_0_ to have any truncated discrete distribution with support on the set {0, 1, 2, …, *N*}, e.g. the Poisson distribution (coined Poisson distribution in the accompanying R-script). Finally, if we know exactly the initial number of infected, we can choose this number to have probability one. The conditional distribution *p*(*y*_0_|*x*_0_) in Equation (3) can be calculated by the approximations outlined in The observational model section.

In summary, we can illustrate the dependency structure of the Hidden Markov model in Figure 6.

**Figure 6:**
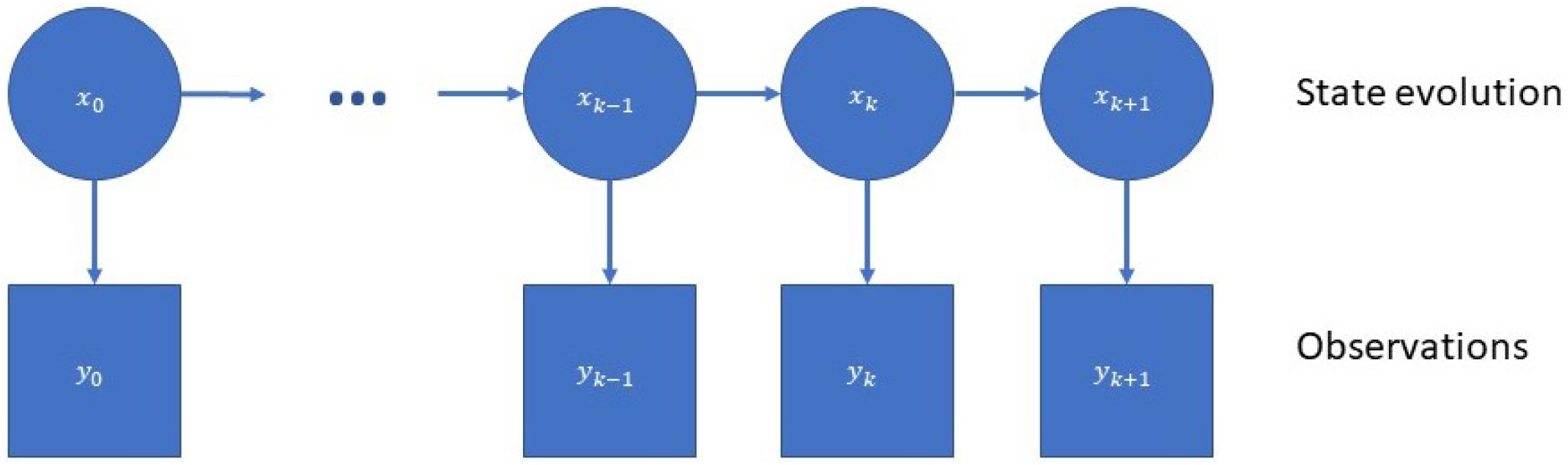
Dependency structure of Bayesian filter

### 4.2 The observation model

Normally, the epidemic model is unobserved, but each day a number of people are tested for infection, and yet a number of the positive samples are sequenced to classify samples into variants. From the sequenced samples, the number of a given variant is recorded. Assuming the number of the PCR and WGS sample sizes, *n*_*k*_ and *m*_*k*_, are known, the sequential sampling scheme can be formulated as a hierarchical model, in the following way:

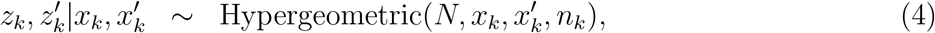

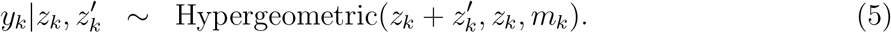

Notice that (4) and (5) are two and one dimensional hypergeometric distributions, respectively. In order to let this be well-defined, we implicitly assume that *y*_*k*_ = 0 if *z*_*k*_ = 0.

Now, it is possible to formulate an expression for the observational model 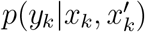, by the following mixture of hypergeometric distributions:

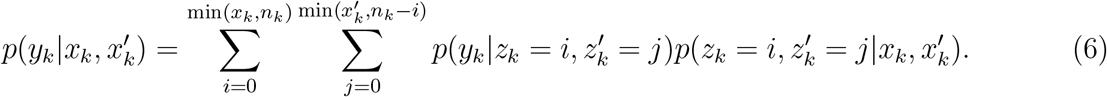

However, due to computational complexity of the involved binomial coefficients, we seek approximations of (6). For the given population sizes and infection rates, we would expect a Poisson approximation to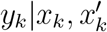, with a mean value matching the ratio of the specific variant in the population, 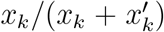, times the sample size, *m*_*k*_, will provide a good approximation of the distribution of 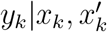, i.e.

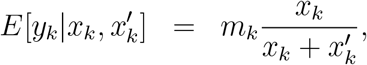

which leads to the following Poisson distribution approximation (Poisson)

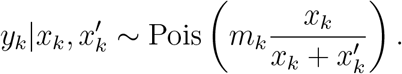

and the following Binomial distribution approximation (Binom1)

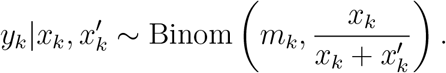

and the following Binomial distribution approximation (Binom2) based on ratios

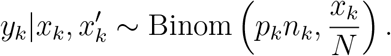

We simulated 10,000 realizations of *y*_*k*_ from the two-stage sampling distrution, with *N* = 600, 000, *x*_*k*_ = 3, 312, 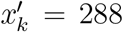, *n*_*k*_ = 17, 000, and *m*_*k*_ = 29, and constructed an approximation to the sampling distribution by the relative frequencies. Next, we compared the Poisson, Binom1, and Binom2 approximations to the approximated sampling distribution by the Kullback-Leibler distance, see Figure 7.

**Figure 7:**
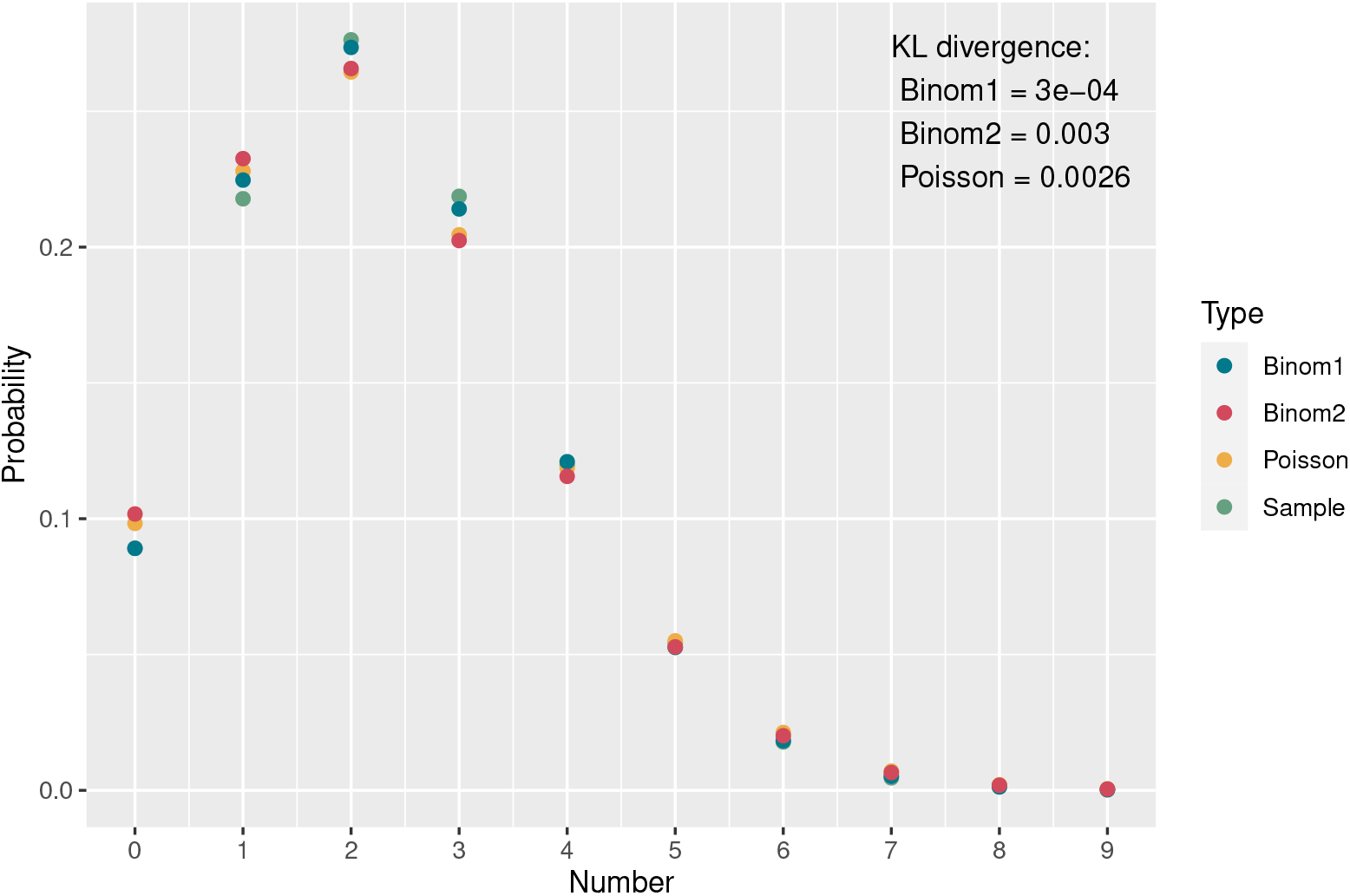
Comparison of approximations and simulated sampling distributions including KulbackLeibler divergence. Parameters used were *N* = 600; 000, 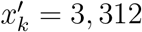, *x*_*k*_ = 288, *n*_*k*_ = 17, 000, and *m*_*k*_ = 29.

To put the use of KL distance for comparison into perspective, consider two Poisson distributions *f*_1_ and *f*_2_ with intensities *λ*_1_ and *λ*_2_ = (1 +)*λ*_1_, where *f*_2_ can viewed as a slight perturbation of *f*_1_. For the two Poisson distributions we have:

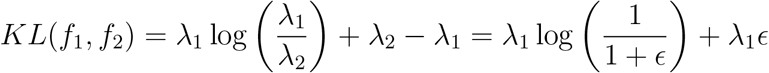

A second order Taylor approximation of *KL* as function yields

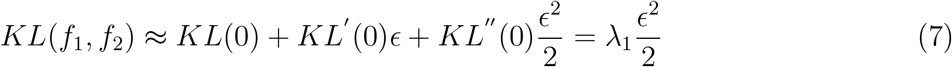

In the simulations above, we have *λ*_1_ = 2.32. If we plug this into (7) together with the simulated KLD, we get = 0.042 and *λ*_2_ = 2.42, illustrating the proximity of the Poisson approximation.

### 4.3 Estimation of the current number of a specific variant

The main question of the paper is to estimate the distribution of the current number of the specific variant given past and current observations of the variant, i.e., the problem is to find *p*(*x*_*k*_|*y*_0_, …, *y*_*k*_).

This can be achieved by a traditional recursive Bayes filter with initial value

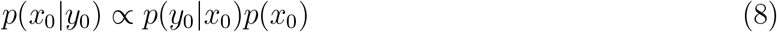

and

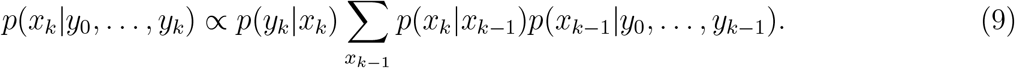

for *k* = 1, 2, 3, …. We notice that, all values for the recursion in (8) and (9) have been specified above in the epidemic model and observation models.

## Data Availability

Data on the weekly number of PCR tests and infected people from Danish Covid-19 test centers are
publicly available from Statens Serum Institut at: https://covid19.ssi.dk/overvagningsdata/
download-fil-med-overvaagningdata
Data on the number of WGS samples per week in the North Denmark region were obtained from
the Danish Covid-19 Genome Consortium at: https://www.covid19genomics.dk/statistics
Data on Danish cluster5 samples were obtained from a dedicated S:Y453F (mink mutation)
build at Nextstrain[4]: https://nextstrain.org/groups/neherlab/ncov/S.Y453F?c=gt-S_453&
f_clade_membership=Mink.Cluster5&f_region=Europe
Population sizes in the municipalities in the North Denmark region were obtained from Statbank,
Statistics Denmark: https://www.dst.dk/en/Statistik/emner/befolkning-og-valg/
befolkning-og-befolkningsfremskrivning

https://covid19vocmonitor.aau.dk

## 5 Data availability

Data on the weekly number of PCR tests and infected people from Danish Covid-19 test centers are publicly available from Statens Serum Institut at: https://covid19.ssi.dk/overvagningsdata/download-fil-med-overvaagningdata

Data on the number of WGS samples per week in the North Denmark region were obtained from the Danish Covid-19 Genome Consortium at: https://www.covid19genomics.dk/statistics

Data on Danish cluster5 samples were obtained from a dedicated S:Y453F (mink mutation) build at Nextstrain[5]: https://nextstrain.org/groups/neherlab/ncov/S.Y453F?c=gt-S_453&f_clade_membership=Mink.Cluster5&f_region=Europe

Population sizes in the municipalities in the North Denmark region were obtained from Stat-bank, Statistics Denmark: https://www.dst.dk/en/Statistik/emner/befolkning-og-valg/befolkning-og-befolkningsfremskrivning

## 6 Code availability

The R code and data are available at https://github.com/HaemAalborg/cluster5. A Shiny app that can be used to run the algorithms is available at https://covid19vocmonitor.aau.dk.

## Acknowledgements

The authors gratefully acknowledge the Novo Nordisk Foundation’s support to the COVID-19-CTRL Project and the Poul Due Jensen Foundation’s support through the BEO-COVID Project.

## Author contributions

The mathematical model was derived by HS, TK, and MB. JS provided input to the mathematical model. Implementation in R and analysis were done by MB and RB. The manuscript was drafted by HS, TK, and MB. All authors read and approved the final manuscript.

