## Supplementary Information for "Mathematical modelling of SARS-CoV-2 variant outbreaks reveals their probability of extinction"

In this appendix a simplified model is presented and analyzed to demonstrate the counter-intuitive non-monotonous relationship between extinction probability and reproduction number. Consider a discrete time random process $x : N \to R_+ \cup 0$ with initial distribution on $\{0, 1\}$. The process constitutes a simple model of the number of infected, and its initial value $x(0) \in \{0, 1\}$ constitutes a simple model of the infection being extinct or not. The evolution of $x$ is defined by $x(n) = \exp(a)x(n-1)$, where $a$ can viewed as a growth rate and $\exp(a)$ is equivalent to the reproduction number. If $x(0) = 0$, then $x(n) = 0$ for all $n \geq 0$, whereas if $x(0) = 1$, then $x(n) = \exp(a)^n = \exp(an)$. At time $N > 0$ a testing procedure is modelled with the assumption, that each individual of (an effectively infinite population) is tested with probability $p$, which means that the probability of finding 0 infected is therefore $(1-p)^{x(N)}$. To model the higher (a-priori) extinction probability of lower growth rates we set the probability of extinction ($P(x(0) = 0)$) as

$$P(x(0) = 0) = \frac{1}{1 + \exp(a/\alpha)},$$

where $\alpha > 0$ is a shaping parameter. Thus

$$\lim_{a \to -\infty} P(x(0) = 0) = 1,$$
$$\lim_{a \to \infty} P(x(0) = 0) = 0.$$

Finally, we can compute the conditional probability $P_{e|0}$ of extinction given 0 infected were found at the test at time $N$ by

$$\begin{aligned} P_{e|0} &= \frac{P(x(0) = 0)}{P(x(0) = 0) + P(x(0) = 1)(1-p)^{x(N)}} \\ &= \frac{\frac{1}{1+\exp(a/\alpha)}}{\frac{1}{1+exp(a/\alpha)} + \frac{\exp(a/\alpha)}{1+\exp(a/\alpha)}(1-p)^{\exp(aN)}} \\ &= \frac{1}{1 + \exp(a/\alpha)(1-p)^{\exp(aN)}} \end{aligned}$$

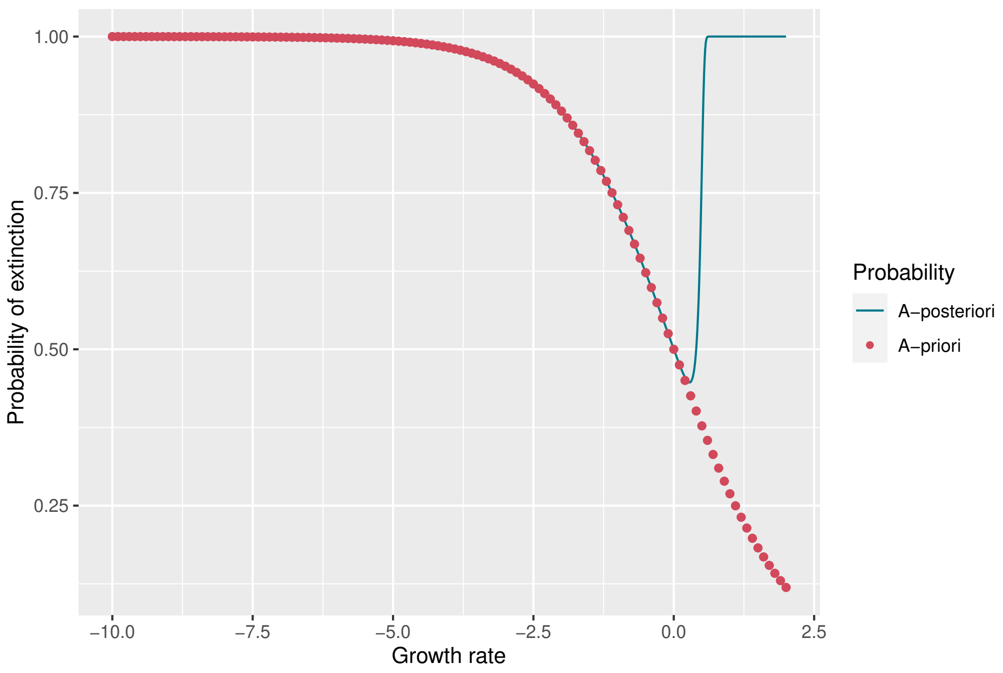


Supplementary Figure 1: The a-priori and a-posteriori extinction probabilities, $P(x(0) = 0)$ and $P_{e|0}$, respectively, for varying growth rates $a$.

The conditional (a-posterior) probability $P_{e|0}$ is shown in Figure 1 for $\alpha = 1, N = 15, p = 10^{-3}$ and varying growth rates. Figure 1 clearly indicates the (counter-intuitive) non-monotonous relationship between growth rate and the a-posterior probability of extinction $P_{e|0}$. $P_{e|0}$ is also minimal for a growth rate slightly higher than 0 as is the case for the elaborate model.
